# Phase I study on the pharmacokinetics of intravaginal, self-administered artesunate vaginal pessaries among women in Kenya

**DOI:** 10.1101/2024.07.08.24309596

**Authors:** Chemtai Mungo, Katherine Sorgi, Brenda Misiko, Cynthia Cheserem, Lisa Rahangdale, George Githongo, Cirilus Ogollah, Jackton Omoto, Mihaela Plesa, William Zamboni

## Abstract

Cervical cancer remains a significant global health issue, especially in low- and middle-income countries (LMICs), where access to prevention and treatment is limited and women are at a higher risk of cervical cancer. Artesunate, a widely available drug used to treat malaria, has shown promise in treating human papillomavirus (HPV)-associated anogenital lesions including high-grade cervical precancer, in a recent Phase I studies in the United States. Data on the pharmacokinetics of artesunate following intravaginal use, and its implications on malaria resistance, are lacking.

**Objectives:** The primary objective of this study is to investigate the pharmacokinetics of Artesunate (AS) and its active metabolite, dihydroartemisinin (DHA) following intravaginal use at the dosing and frequency intended for cervical precancer treatment. A secondary objective is to assess safety among study participants.

**Methods:** We are conducting a single-arm, phase I trial with a sample size of 12 female volunteers. Participants will self-administer artesunate vaginal pessaries in the study clinic daily for 5 consecutive days. Participants will have their blood drawn prior to receiving the first dose of artesunate on day one of the study and then will receive 8 blood draws on study day five, prior to artesunate administration and at 15 minutes, 30 minutes, 1 hour, 2 hours, 4 hours, 6 hours, and 8 hours after pessary administration. Pharmacokinetic parameters of artesunate and DHA will be calculated by way of quantitative analysis of with determination of maximum concentration (Cmax), time to Cmax (Tmax), area under the serum concentration versus time curve (AUC), apparent clearance, and elimination half-life (t1/2).

## Introduction

Cervical cancer is the second most common cancer in women globally, with a high prevalence in low- and middle-income countries like Kenya. It is caused by human papillomavirus (HPV) infection but can be prevented through vaccinations or early screening to find precancerous changes in the cervix – also known as cervical intraepithelial neoplasia. While these methods can prevent cervical cancer from developing in women, there are access and resource barriers, especially in LMICs. For instance, HPV vaccinations are new in Kenya and other LMICs, and very few women under the age of 20 have been vaccinated. There are also cost barriers and shortages in healthcare providers. In countries like Kenya and other LMICs where there are few nurses and doctors, many women with cervical precancer are often referred to far away facilities to access treatment and due to costs and other challenges, many are unable to access the referral centers with providers who can offer treatment.

These barriers have led to researchers exploring the option of self-administered treatments for cervical precancer to overcome healthcare provider and healthcare resource shortages. One treatment being studied is an intravaginal pessary formulation of the drug Artesunate, which is often used to treat malaria. While a U.S. study has shown its safety and early efficacy for cervical precancer treatment,^1^ it is unknown whether intravaginal use of artesunate vaginal inserts at the dosing and frequency used, may result in systemic exposure and hence have implications for malaria resistance. To fill this gap in the literature, this study seeks to investigate the pharmacokinetics of intravaginal artesunate at the dose and frequency used in ongoing cervical precancer treatment studies.

### Intravaginal Artesunate for Cervical Precancer Treatment

Artesunate (AS), a World Health Organization (WHO) approved malaria drug, is being explored as a potential topical therapy for cervical precancer as its safety and tolerability has long been well established.^2–5^ Growing evidence suggests its cytotoxic effects against numerous cancer cell lines both *in vitro* and *in vivo* and have revealed its proposed mechanisms of action include suppressing cell proliferation by inducing G1 and G2M phrase cell cycle arrest and modulation of inflammatory pathways characteristic of uncontrolled proliferation and carcinogenesis.^6–9^ Ferroptosis, an iron-dependent cell death type, is thought to be a key anticancer mechanism for HPV-infected cells.^10^ Cancer cells are highly proliferative, requiring a heavy iron load which acts as a cofactor in synthesizing deoxyriboses before cell division.^11^ Development of both high-grade cervical intraepithelial neoplasia (CIN2/3), the precursor lesion of cervical cancer, and cervical cancer are associated with the expression of two viral proteins in the HPV lifecycle, E6 and E7.^12^ Epithelial cells that express either or both of these oncoproteins also overexpress the transferrin receptor, and have been shown to have increased levels of intracellular iron compared with normal cells.^10^ This observation has been exploited to investigate whether preinvasive cervical cancer (CIN2/3), can be treated with Artesunate, which contains an endoperoxide bridge that reacts with intracellular ferrous iron to generate free radicals, capable of inducing direct oxidative damage resulting in cell death.^9^ Given the mechanism of action, artesunate may provide beneficial and therapeutic effects for intraepithelial HPV disease.

Several studies demonstrate the pro-apoptotic effects of Artesunate by activating the mitochondria-dependent pathway, including caspase-3/9 activation and cytochrome c release from the mitochondria.^13^ It induces autophagy in uterine corpus endometrial carcinoma cells and elevates reactive oxygen species in human bladder cancer cells and hepatocellular carcinoma cells.^14–16^ Artesunate has demonstrated anti-angiogenic properties by downregulation VEGF and angiopoietin-1 in myeloma cells.^17–19^ Its reported anti-cancer effects involve the formation of alkaline radicals through an endoperoxide bridge, reacting with intracellular ferrous iron and leading to cell death.^20^ The overexpression of the transferrin receptor in cervical squamous cell cancers and their precursors,^21^ cervical intraepithelial neoplasia (CIN), prompted a study of cytotoxic effects of dihydroartemisinin (DHA), the bioactive form of Artesunate, on papillomavirus-expressing epithelial cells.^22^ *In vitro* studies revealed that DHA had minimal impact on normal cervical epithelial cells but had significantly induced cytotoxicity in HPV-immortalized cervical cells.^22^ Administered as a local treatment in a canine nonclinical model with a 100% known tumor growth rate with HPV-infection, DHA (2.22 mg dissolved in 100 µl dimethyl sulfoxide) reported to inhibit papillomavirus-induced tumor formation. In addition, tumor-negative dogs developed antibodies against the HPV L1 capsid protein.^22^

### Safety for Intravaginal Use

In a multi-center dose-escalation phase 1 study in the United States, intravaginal artesunate inserts (pessaries) were tested for safety, tolerability, and efficacy in women with CIN2/3.^1^ This “first-in-human” study demonstrated that intravaginal Artesunate for CIN2/3 treatment was safe, well tolerated, and resulted in self-limited adverse events that were graded I and II. Reported AEs among participants who used 3 five-day artesunate cycles included chills and flu-like symptoms (n=3, grade 1), vaginal (yeast) infection (n=1, grade II), dizziness or headache (n=2, grade 1), non-infective cystitis (n=1, grade 2), vaginal pain or uterine cramping (n=9, grade I), vaginal discharge (n=4, grade 1), vaginal pruritis (n=9, grade 1). In summary, 37 drug-related AEs were observed in this Phase I trial, of which 34 (92%) were grade I, and 3 (8%) were grade 2. Reported grade 2 adverse events included vaginal yeast infection (n = 6), bacterial vaginosis (n = 2), vaginal inflammation (n = 2), urinary tract infection (n = 2), and noninfective cystitis (n = 1). Adverse events that were determined to be unrelated to the study medication included: anxiety, insomnia, suicidal ideation, vaginal twitching, fever, flu-like symptoms in a patient who developed a cold, body itching, chills, and eczema flare. No subjects withdrew from the study because of intolerable side effects, and all 28 subjects included in the modified-intention-to-treat analyses were able to complete their designated dosing regimen. No grade 3 or 4 adverse events were reported, and three subjects reported no noticeable side effects. Currently, a randomized placebo-controlled phase II study of artesunate vaginal inserts in treatment of CIN2/3 is enrolling subjects at several US sites, with no serious adverse events reported so far (NCT04098744). Additionally, a phase II randomized trial of artesunate suppositories for treating anal high-grade squamous intraepithelial lesions among HIV-negative individuals is ongoing in the United States (NCT5555862)

### Summary of Artesunate Pharmacokinetics

The pharmacokinetics (PK) measures of artesunate (AS) and dihydroartemisinin (DHA) following intravenous (IV), intramuscular (IM), oral and rectal administration have been well described.^23^ These PK parameters include maximum concentration of the drug in the blood (Cmax), time to maximum concentration Cmax (Tmax), apparent clearance (CL/F) - the drug concentration in the body in proportion to the rate of elimination, volume of distribution (V/F), the area under the serum concentration versus time curve (AUC) which expresses the total amount of the drug in systemic circulation after administration, and half-life (t_1/2_). IV administration of AS quickly produces high maximum concentrations (Cmax) of AS, higher than any other method of administration. One example of this can be seen in a study of adults with uncomplicated malaria which compared AS and DHA levels following IV and IM administration, Cmax values for AS when administered intravenously reached over 16,000ng/mL while only reaching around 884mg/mL when administered intramuscularly.^24^ Similarly, the maximum concentration (Cmax) of AS and DHA peak quickly following IV administration, followed by a rapid decline. The average half-life of AS following IV administration of less than fifteen minutes in multiple studies, with an observed clearance range of 2-3 L/kg/hr and a volume range of 0.1-0.3 L/kg.^23^ The hydrolysis of AS into DHA following IV administration is similarly quick, with maximum DHA levels reached soon after IV AS administration. The Tmax for DHA after IV administration was consistently less than 25 minutes according to the observed studies with DHA clearance averaging between 0.5-1.5 L/kg/hr and volume averaging between 0.5-1.0 L/kg.^23^ The AUC of AS following IV administration of 120 mg AS in adults ranged between 876ng*hr/ml in healthy volunteers^25^ and 1038 - 1269ng*hr/ml in those with malaria.^26^ The AUC of DHA following IV administration of 120 mg AS in adults ranged from 1850 ng*hr/ml in healthy volunteers^27^ to 1845-2377ng*hr/ml in those with malaria.^28^

Compared to IV administration, IM administration produces lower peaks, longer half-life, and higher volumes of distribution for AS, as well as delayed peaks for DHA. For example, AS half-life following in IV administration is less than 15 minutes on average, compared to 25.2 to 48.2 minutes with IM administration.^23^ Other parameters – in including DHA half-life, volume of distribution, and clearance rates following IM administration resembled the values recorded after IV administration in multiple studies, due to the high bioavailability, assessed by exposure to DHA, associated with IM AS administration (>86%).^23^ The AUC of AS following IM administration of 120 mg AS in adults ranged between 856^29^ - 999^25^ng*hr/ml in those with malaria. The AUC of DHA following IM administration of 120 mg of AS in adults with malaria was 2474ng*hr/ml in one study.^25^

When AS is administered orally, DHA peak concentrations (Cmax), AUC, and half-life averages are all notably higher than comparable AS parameters. The average time to DHA Cmax and half-life following oral AS administration (200 mg/day) are 2 hours, and 0.5-1.5 hours, respectively, compared to one hour and 20-45 minutes, respectively, for AS.^23^ While a similar pattern is seen following IV and IM administration, namely, elevated levels of DHA half-life and AUC compared to AS, the difference are notable following oral administration. Also, while oral administration of AS results in a higher DHA Cmax compared to AS, IV and IM administration result in a notably higher AS Cmax than DHA Cmax. Morris et al (2011) points out that the variations observed following oral administration are most likely attributed to AS functioning as a “pro-drug” for DHA when ingested orally and in response to “first-pass or systemic metabolism.”^23^ Essentially, when AS is taken orally, it is converted to DHA at a greater extent than when it is taken intravenously or intramuscularly. The AUC of AS and DHA in a study of healthy adult volunteers taking 200mg AS daily for 5 days was 67 ng*hr/ml and 1158 ng*hr/ml for AS & DHA, respectively, on Day 1, and 60 ng*hr/ml and 1300 ng*hr/ml for AS & DHA, respectively, on Day 5.^30^ Similar Cmax parameters for AS were 67 ng/ml and 58 ng/ml on Day 1 and 5, respectively, and a pooled DHA Cmax of 654 ng/ml, demonstrating the absence of time-dependent artesunate pharmacokinetics in healthy subjects during 5-day oral administration of 200 mg artesunate.^30^ The AUC of AS following oral administration of 200 mg AS in adults ranged between 60-67ng*hr/ml in healthy volunteers^31^ and 310ng*hr/ml in those with malaria.^32^ The AUC of DHA following oral administration of 200 mg of AS in adults ranged from 1158-1331 ng*hr/ml^31,33^ in healthy volunteers to 3027ng*hr/ml in those with malaria.^32^ Rectal AS administration yields pharmacokinetic results similar to those obtained from oral administration, with the exception of delayed AS Cmax and longer AS half-life. Compared to IV administration, expectedly, both AS absorption and elimination are prolonged following rectal administration. Following rectal administration of AS, Tmax average between 0.58-1.43 hours, with a half-life between 0.9-.95 hours.^23^ These averages are based on three different studies: two studies containing pediatric patients with uncomplicated falciparum malaria (10 - 20mg/Kg dosing)^34,35^ and one study containing healthy Malaysian adults (200 mg rectal suppository, ∼4mg/Kg dosing).^33^ Following rectal dosing of a one-time 200 mg AS suppository in healthy adults (similar to our planned dosing), a Cmax, Tmax, half-life and AUC of 448.5 ng/ml, 1.43 hours, 0.95 hours, and 796 ng*hr/ml of AS were observed, and Cmax, Tmax, half-life and AUC of 385.6 ng/ml, 1.80 hours, 1.21 hours, and 965 ng*hr/ml respectively of DHA were observed in healthy adults.^33^ No data are available on rectal PK in adults with malaria as this route of administration is not used to treat malaria in adults. The longer half-life of AS following rectal dosing (average 0.9-0.95 hrs) compared to IM (average 25.2 – 48.2 minutes), or IV (average less than 5 minutes) may reflect absorption rate-limited elimination of AS.^23^ As is expected given that rectal AS administration avoids by-pass metabolism, the discrepancy in AS and DHA AUC values (796 ng*hr/ml and 965 ng*hr/ml, respectively), is not as striking with rectal, as compared with oral administration of AS (119 ng*hr/ml and 1331 ng*hr/ml, respectively).^23^ Similar to oral administration, both DHA Tmax and half-life values were higher than that of AS following rectal administration.^23^

### Effect of malaria infection status on artesunate and DHA pharmacokinetics

Teja-Isavadharm et al., conducted a direct comparison of DHA pharmacokinetics following oral AS administration in healthy adults and falciparum malaria patients.^36^ The investigators found significantly higher AUC and Cmax of DHA in subjects with malaria as compared to healthy subjects. Similar results were obtained by Binh et al in a study comparing the PK in eight patients with falciparum malaria and ten healthy subjects.^37^ Due to the small size of both studies, definitive conclusion regarding differences in PK between healthy and infected subjects cannot be drawn.^27^ However, as DHA clearance is dependent on hepatic blood flow, a reduction in clearance, and consequently an increase in exposure associated with acute malaria infection, would be consistent with known DHA’s PK properties.^27^

### Malaria treatment and artemisinin resistance

Artemisinin-based combination therapies (ACTs) are recommended by the WHO as the first line treatment for uncomplicated Plasmodium Falciparum.^38^ In ACTs, artemisinin quickly, and drastically, reduces the majority of malaria parasites, with the partner drug clearing the remaining parasites to prevent recrudescence.^39–40^ Artemisinin resistance is defined as delayed parasite clearance (following treatment with an artesunate monotherapy or ACT) observed as a parasite clearance half-life greater than five hours or microscopic evidence of parasites on day three.^41^ This represents partial resistance. While artemisinin resistance alone does not necessarily lead to malaria treatment failure, reduced efficacy of the artemisinin component places greater demands on the partner drug to clear a larger parasite mass, jeopardizing future efficacy (WHO). Examples of ACTs include artemether-lumefantrine, artesunate-amodiaquine, artesunate-mefloquine, among others. Standard ACTs regimen for uncomplicated malaria is an oral 3-day course. Most studies indicate that current ACTs recommended in national malaria treatment policies remain effective, with an overall efficacy rate of greater than 95%.^42^

Artemisinin’s act exceptionally fast against intra-erythrocytic asexual blood-stage malaria parasites, affecting up to 10,000-fold reductions in parasite burden every 48 hours.^43^ The primary genetic drivers of artemisinin resistance, both *in vitro* and *in vivo,* are point mutations in the P. falciparum Kelch13 (*PfK13*) gene during the early ring stage.^44,45^ These mutations allow a subset of early ring-stage parasites to survive cell-cycle arrest brought on by artemisinin exposure, enabling those parasites to reinitiate transcription and complete their intraerythrocytic developmental cycle once artemisinin is no longer present at inhibitory concentrations.^46^ *In vitro* resistance is routinely defined as greater than 1% survival of early ring-stage parasites exposed to 700nM dihydroartemisinin (DHA-the primary active metabolite of ART) for 6 hours, followed by drug-free culture incubation for a further 66 hours.^41^ The resistance mechanism appears to involve a complex interplay of K13 protein abundance, hemoglobin endocytosis, and the parasite response to stress.^41,47^

In Africa, several studies have identified a number of low-frequency Pfk13 mutations associated with delayed parasite clearance in four countries: Ghana, Rwanda, Uganda Tanzania.^48,49^ Mutations including M476I, P553L, R561H, P574L, C580Y and A675V, were observed at low frequencies under 5%.^48^ For example, in Tanzania, mutations were found in two parasites from 764 samples in 2027 (0.3%)^6^, and one parasite from 422 samples in 2019 (0.2%).^48^ Similarly in Uganda, one parasite was identified from 796 samples in 2018/2019 (0.1%).^50^ A 2021 study in Northern Uganda from 2017 to 2019 identified in vivo artemisinin resistance (parasite clearance half-life >5 hours) in a total of 14 out of 240 patients who received intravenous artesunate.^50^ Of these 14 patients, 13 were infected with P. falciparum parasites with mutations in the A675V or C469Y allele in the kelch13 gene.^45^

P. falciparum resistance to artemisinin has been documented in five countries in Southeast Asia; Cambodia, Lao People’s Democratic Republic, Myanmar, Thailand and Vietnam (WHO).^40^ With implementation of combination therapy, improvements to health systems and surveillance systems to monitor first- and second-line treatment, the consequences of the development of resistance to antimalarial medicines may be less severe today than what was observed with chloroquine in the 1980s. If parasites develop reduced sensitivity to artemisinin, ACTS will continue to cure malaria, as long as the partner drug remains effective.^40^ To overcome resistance, potential changes can be made to ACT. Some of these include modifications such as extending the duration of the ACT course (currently 3 days for oral treatment), alternating use of different ACT regimens, and addition of another antimalarial drug to the standard ACTs (triple-ACT).^40^ Additionally, adding a malaria vaccine (e.g. RTS, S vaccine) to mass drug administration campaigns could enhance treatment efficacy and help prevent further artemisinin resistance development.

### Systemic Artesunate absorption and possible implications for developing resistance for malaria treatment

Although there is no available pharmacologic data on serum absorption following intravaginal artesunate administration, this study aims to address this gap. The direct application of artesunate to the cervical mucosa at the proposed dose of 200mg (≤4 mg/kg based on planned inclusion criteria of weight≥ 50 Kg) is unlikely to result in systemic absorption. This planned dosage is 2.5-fold lower than the approved rectal suppository dose (10mg/Kg). In contrast to the rectal mucosa, which is highly vascular and comprised of single-cell layer of columnar epithelium making it highly permeable, the cervico-vaginal tissue has a thick, stratified squamous epithelial cell layer and is significantly less vascular, reducing its systemic absorption.^51^ Similarly, due to the rapid rate of elimination of Artesunate’s active metabolite,^52^ no systemic accumulation is expected of Artesunate or its active metabolite with intravaginal multi-day dosing in the context of cervicovaginal administration.

## Methods

### Study Objectives Primary Objective

The primary objective is to determine the area under the serum concentration versus the time curve (AUC) of DHA following five consecutive days of self-administration of Artesunate vaginal inserts among healthy women living in Kenya

### Secondary Objectives

1. Determine the area under the serum concentration versus time curve (AUC) of Artesunate (AS) following five consecutive days of self-administration of 200mg Artesunate vaginal inserts (pessaries) among healthy women in Kenya
2. Determine the maximum concentration of Artesunate (AS) (Cmax) following five consecutive days of self-administration of 200mg Artesunate vaginal inserts (pessaries) among healthy women in Kenya
3. Determine the maximum concentration of dihydroartemisinin (DHA) (Cmax) following five consecutive days of self-administration of 200mg Artesunate vaginal inserts (pessaries) among healthy women in Kenya
4. Determine the time to maximum concentration (Tmax) of Artesunate (AS) following five consecutive days of self-administration of 200mg Artesunate vaginal inserts (pessaries) among healthy women in Kenya
5. Determine the time to maximum concentration (Tmax) of dihydroartemisinin (DHA) following five consecutive days of self-administration of 200mg Artesunate vaginal inserts (pessaries) among healthy women in Kenya
6. Determine the half-life (t_1/2_) of Artesunate (AS) following five consecutive days of self-administration of 200mg Artesunate vaginal inserts (pessaries) among healthy women in Kenya
7. Determine the half-life (t_1/2_) of dihydroartemisinin (DHA) following five consecutive days of self-administration of 200mg Artesunate vaginal inserts (pessaries) among healthy women in Kenya
8. Determine the apparent clearance (CL/F) of Artesunate (AS) following five consecutive days of self-administration of 200mg Artesunate vaginal inserts (pessaries) among healthy women in Kenya
9. Determine the apparent clearance (CL/F) of dihydroartemisinin (DHA) following five consecutive days of self-administration of 200mg Artesunate vaginal inserts (pessaries) among healthy women in Kenya
10. Determine the volume of distribution (V/F) of Artesunate (AS) following five consecutive days of self-administration of 200mg Artesunate vaginal inserts (pessaries) among healthy women in Kenya
11. Determine the volume of distribution (V/F) of dihydroartemisinin (DHA) following five consecutive days of self-administration of 200mg Artesunate vaginal inserts (pessaries) among healthy women in Kenya
12. Investigate the safety of 5-day course of self-administered intravaginal artesunate vaginal inserts (pessary) in women in Kenya

### Study design

This is a single-arm, nonrandomized, interventional phase I study among 12 women over the age of 18 living in Kisumu, Kenya. Participants will self-administer 200 mg artesunate vaginal inserts for five consecutive days with blood draws performed on day 1 (at baseline) and on day 5. Day 1 will consist of one blood draw prior to first pessary use and day 5 will consist of eight blood draws: one prior to final artesunate administration, at time 0, then at 15 minutes, 30 minutes, 1 hour, 2 hours, 4 hours, 6 hours, and 8 hours after final artesunate pessary insertion. The schedule of events is outlined in Figure 1.

### Study setting

This study will be conducted in Kisumu, Kenya at the Lumumba Sub-County Hospital and at the Victoria Biomedical Research Institute. All study activities will take place at Lumumba Sub-County Hospital except the day 5 study visit, which will take place at Victoria Biomedical Research Institute.

### Study Drug

In this study, a vaginal insert (pessary) formulation of artesunate will be used. Artesunate is a derivative of artemisinin which is the active principle of the Chinese medicinal herb *Artemisia annua.* The quantitative composition of artesunate inserts utilized in this study is detailed in Table 1.

**Table 1:** Composition of artesunate inserts.

| Ingredient | Function |
| --- | --- |
| Artesunate | Active ingredient |
| PCCA MBK<sup>TM*</sup> - hydrogenated vegetable oil and PEG-8 distearate. | Base |
| Butylated Hydroxyl Anisole NF | Preservative |

The drug is produced by Buderer Drug Co in Perrysburg Ohio and is compounded in accordance with USP Chapter 795 (Pharmaceutical Compounding – Nonsterile Preparations) and USP Chapter 797 (Pharmaceutical Compounding – Sterile Preparations). These chapters establish practice standards and outline the responsibility of the compounder, selection and appropriate sources of ingredients, quality control, and considerations regarding the stability of compounded preparations. Additionally, USP Chapter 1075 (Good Compounding Practices) and USP Chapter 1160 (Pharmaceutical Calculations in Prescription Compounding) provide further guidance ensuring the quality and accuracy of the compounded Artesunate.

For each compounded lot of vaginal inserts used in this study, potency testing is conducted to confirm the dosage of Artesunate. Each lot has an expiration date of 6 months after compounding, although testing on batches beyond this period has demonstrated no significant loss in strength. This formulation, at various doses, has been used in two Phase 1/2A clinical trials for cervical and anal HSIL^1,53^.

### Eligibility and Recruitment

#### Recruitment

Participants will be recruited from the general population within close proximity to the study location in Kisumu County, which will include recruitment from local health facilities. The study team will conduct local outreach activities and educational talks in the community to harness interest in participating. If an individual is interested in participating, they will be screened for eligibility and, if eligible, will provide their informed consent and will be briefed on other study procedures before any study activities are performed. Inclusion and exclusion criteria for this study is outlined in Table 2. Recruitment started on 29/05/2024.

**Table 2:**
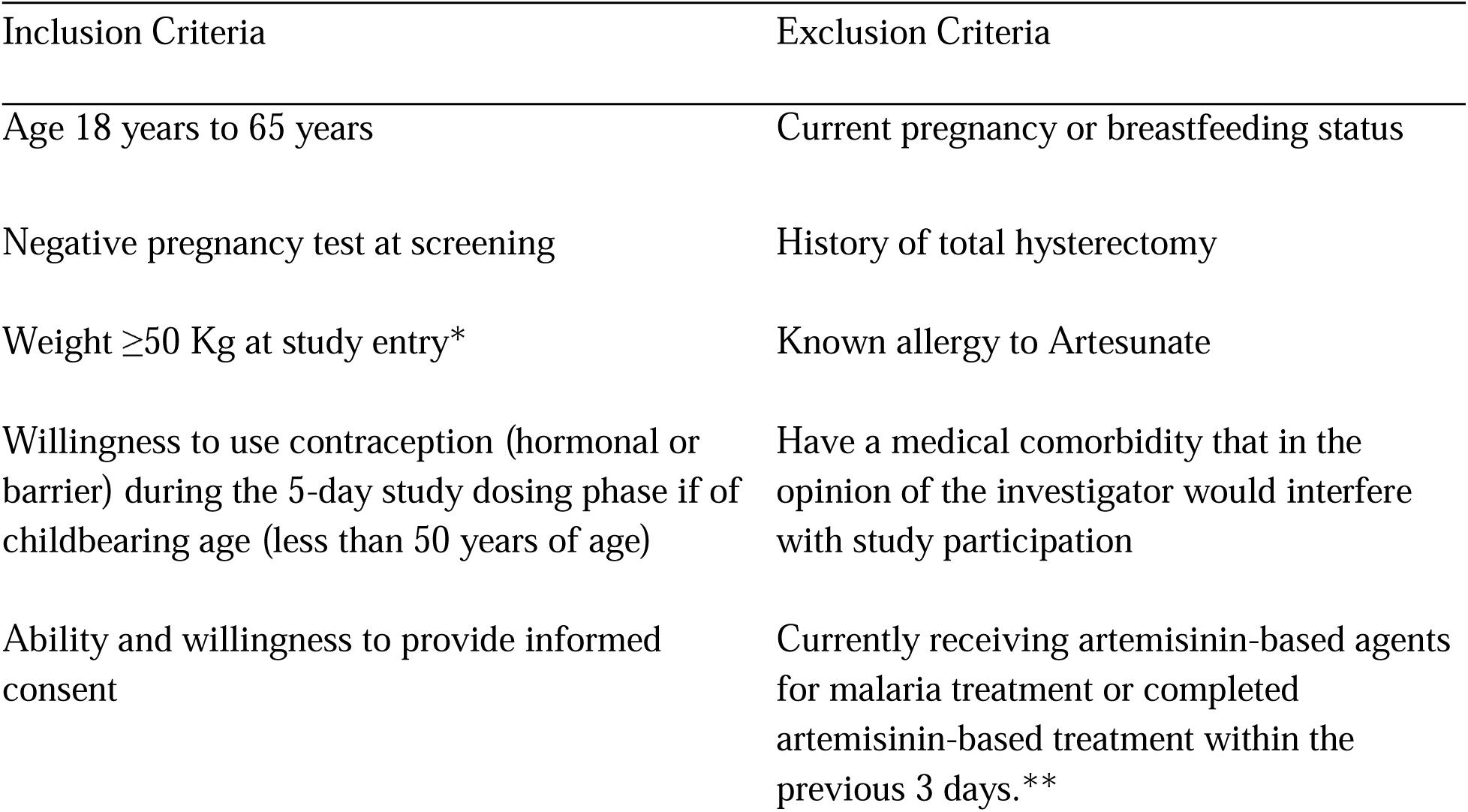
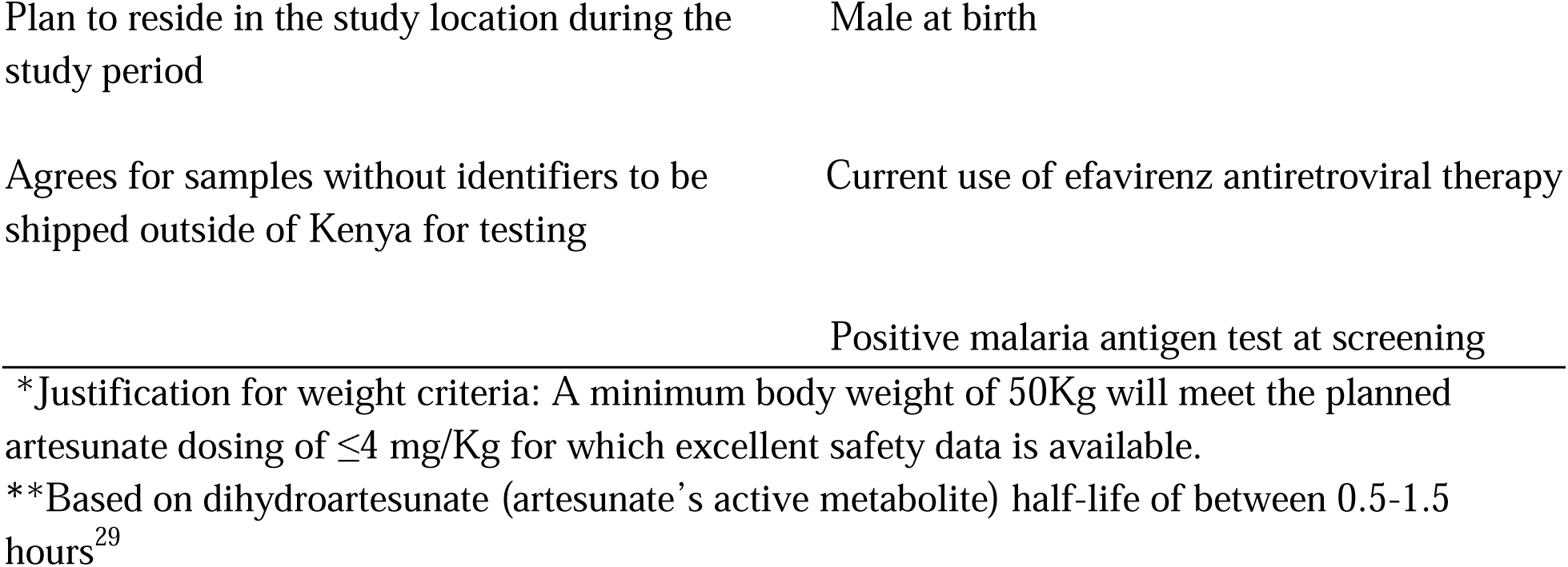
Inclusion and Exclusion Criteria.

#### Study procedures by visit

Pre-screening visit: Prior to enrollment of a participant, the study staff will pre-screen potential participants at on-site or off-site locations. During these visits, study staff will explain the aspects of the study to potential participants and will provide an explanation of eligibility requirements.

Screening/Enrollment visit: To begin the screening process, eligible participants will provide their written informed consent for the study procedures. Once informed consent is obtained by the study staff, the screening processes will begin, including a malaria antigen test to screen for subacute malaria infection. If a participant tests positive for malaria, they will be referred for treatment and considered screen failures, resulting in their discontinuation from participating in the study. Given that the study is located in a malaria-endemic region, we have planned for a high screening-to-enrollment ratio due to the likelihood of high rates of sub-clinical malaria. Other screening and enrollment activities will involve the collection of basic demographic and clinical data, study protocol training (including demonstration of intravaginal artesunate application using a pelvic model), a limited physical and pelvic exam, a urine pregnancy test, and a review of prohibited medications.

Visit 1: On the same day or up to 14 days after the screening and enrollment visit, the participant will have their first study visit in the clinic. During this visit, eligibility will be confirmed, the study protocol will be reviewed, and study staff will provide instructions on self-administering artesunate. Additionally, the participant will have 2.5ml of blood drawn prior to their first intravaginal self-administration of artesunate. Once self-administration of artesunate is complete, the participant will insert a tampon and will be observed for 30 minutes prior to scheduling their second visit on the following day.

Visits 2-4: Visits 2, 3, and 4 must occur within 24 hours of each other so the study staff will attempt to schedule the visits at approximately the same time each day. During these visits, the study staff will review and record any adverse events (AE) using a standardized questionnaire and the U.S. National Cancer Institute Common Terminology Criteria for Adverse Events (CTCAE). If an adverse event is scored a grade II or worse, a pelvic exam will be performed. After review of adverse events, and reiteration of prohibited medications, the participant will be asked to again self-administer artesunate followed by insertion of a tampon. Once they are observed for 30 minutes, the following visit will be scheduled.

Visit 5: Visit 5 will occur no more than 24 hours after visit 4, with the study staff aiming to schedule it at approximately the same time as the prior day’s visit. During this visit, the study staff will review and record adverse events, conducting a pelvic exam as needed based on the grading of observed AEs. After AEs are reviewed and a reiteration of prohibited medications is given, the participant will undergo their first 2.5ml blood draw of the visit before the insertion of artesunate. A peripheral cannula will be inserted at this time for subsequent blood draws.

Following the initial blood draw, the participant will perform their final self-administration of artesunate, followed by the insertion of a tampon. Once the final self-administration of artesunate is complete, 2.5ml of blood will be drawn after 15 minutes, 30 minutes, 1 hour, 2 hours, 4 hours, 6 hours, and 8 hours. After the last blood draw, participants will be scheduled for the final visit, which will take place 4-10 days after visit 5.

Visit 6: Visit 6 will occur 4-10 days after visit 5. During this visit, the study staff will review and record adverse events since the last visit. Once AEs are recorded, the study staff will initiate study termination and will provide the participant with any necessary financial reimbursement.

#### Participant retention plan

Prior to any study procedures, the informed consent document will be reviewed with participants to ensure a clear understanding of the study purpose, procedures, duration, compensation, and any other factors that are necessary to understand. To document their willingness to participate, each participant and a study personnel will sign the informed consent documents. Once enrolled and consented, the study staff will consistently follow-up with participants to ensure they attend all scheduled appointments, to report any adverse event experiences, and to answer any questions that the participant may have about their role in the study. To compensate for their time, each participant will be reimbursed Kshs 1000 (approximately $10) for each visit to account for potential loss of wages and will be reimbursed for transport to and from the clinic. For the final visit, which is expected to last up to 8 hours, participants will be reimbursed an additional Kshs 2,500 ($25) for loss of wages. Participants may also receive refreshments during the visit depending on how long they are asked to wait. If there is ever concern that a participant is lost to follow-up, the study staff will make every effort to regain contact with the participant, whether that be through calls or home visits.

### Statistical consideration

#### Sample size

This study will have a sample size of 12 healthy volunteers.

#### Sample size justification

Participants will self-administer 200 mg artesunate vaginal inserts for five consecutive days. Serum PK studies will be performed in 12 patients with blood draws performed on day 1 (single sample at baseline) and day 5 (serial samples). As this is the first PK study evaluating the serum PK after intravaginal administration of artesunate, there are no directly comparable studies to guide sample size determination. However, evaluating the PK of artesunate at the same dose (200mg) in 12 participants is a relatively high number as most studies evaluate the PK of drugs in n=3 or n=6 participants per dose.^54–56^ In addition, other PK studies of oral or intramuscular artesunate in this population have been performed with approximately 12 patients.^33, 57–59^ This, we expect our sample size of 12 to be sufficient to meet our primary objective. If the PK variability of artesunate after intravaginal administration is greater than anticipated and cannot be adequately evaluated in 12 participants, then the protocol can be amended to enroll an additional n=3-6 participants.

#### Data analysis

The data analysis plan for this study involves assessing the primary outcome, which is the serum concentration versus time curve (AUC) of dihydroartemisinin following five days of self-administration of 200mg Artesunate pessaries. With the primary endpoint being Mean DHA AUC (ng*hr/ml) with standard deviation on Day 5, statistical analysis will compare this mean DHA AUC to historical studies after intravenous (IV), oral, and rectal administration among adults with similar dosing. Results will be analyzed using a student t-test for one-sample observation and a p-value of 0.05 will be considered statistically significant.

The secondary outcomes include assessing the AUC of Artesunate, maximum concentration of Artesunate and DHA (Cmax), time to reach maximum concentration (Tmax) for Artesunate and DHA, half-life (t1/2) for Artesunate and DHA, apparent clearance of Artesunate and DHA, and the volume of distribution of Artesunate and DHA. Statistical analysis of these variables will include comparing data results to historical studies using relevant test materials. The final outcome to be evaluated is the safety of the 5-day self-administration of Artesunate vaginal inserts. This will be done by monitoring and reporting adverse events and categorizing by their severity, with grade 3 or higher being considered severe. The proportion of participants with a severe AE will be reported along the exact (Clopper-Pearson) one-sided upper 95% confidence bounds. Safety among participants will be monitored and reported starting at the first dose of artesunate.

This study will determine conventional pharmacokinetic parameters for Artesunate and dihydroartemisinin (DHA). Parameters such as AUC, Cmax. Tmax, half-life, apparent clearance, and the volume of distribution will be calculated using non compartmental analysis from the serum concentration-time data. AUC for Artesunate will be determined by linear trapezoidal summation with extrapolation to infinity, starting from drug administration to the last observation. All parameters will be calculated using time in hours after the first drug administration. With respect to DHA, AUC will be calculated to the last drug measurable time point. The elimination rate constant (beta) will be calculated from the slope of the terminal phase of the log concentration-time profile, and the elimination half-life (t1/2) calculated from the ratio of ln 2/beta. Other PK parameters will be calculated using standard model-independent formulae.^38^ The estimates of PK parameters for DHA will assume complete conversion of AS to DHA as reported previously.^35^ Data will be plotted graphically and analyzed using Stata 18 software.^60^ Data will be presented as mean with standard deviation (SD).

#### Quality Control Standards

The sample processing quality control standards are described in Table 3 below.

**Table 3:** Laboratory Quality Control Standards.

| Sections | Details |
| --- | --- |
| **Quality standards** | Criteria applied to samples & tests<br>- Analytic methods are validated according to the latest Food and Drug Administration (FDA) bioanalytical method validation guidance<sup>61</sup><br>- Calibration curve with at least 6 non-zero standards, processed in duplicates per run<br>- Calibration standards at lower and upper limits must be included<br>- Each run includes a double blank, blank sample, and 3 quality control samples (low, medium, high), processed in triplicates |
| **Acceptable limits** | Acceptable ranges for tests<sup>62</sup><br>- For calibration curve use in quantification, at least 6 non-zero standard samples must be within +/- 15% nominal concentration, except lower limit of quantification (LLOQ), which must be within +/- 20%<br>- Quality Control (QC) standards acceptance criteria: +/- 15% of nominal value, with at least 67% passing at low, medium, and high QC levels |
| **Analysis methods** | Assay details & limits of quantification for Artesunate and DHA<sup>63</sup><br>- Liquid chromatography-mass spectrometry (LC-MS/MS) |
|  | method will be used for detection and quantification<br>- Simultaneous measurement of artesunate and DHA from the same sample aliquot<br>- LLOQ: 5 ng/ml for artesunate, 8 ng/ml for DHA |
| **Quality control** | Quality control (QC) procedures, acceptable ranges, and batch acceptance criteria<br>- Each batch includes calibration standards and 3 QC samples meeting acceptance criteria outlined above<br>- LLOQ must have a signal-to-noise ratio $\geq 5$<br>- Low QC: 3xLLOQ<br>- Medium QC: 50% of upper limit of quantification (ULOQ)<br>- High QC: 80% ULOQ |

#### Ethics approval and consent to participate

This clinical trial has full ethics review board approval from the University of North Carolina Chapel Hill and the African Medical Research Foundation. Written informed consent will be obtained from all study participants.

## Discussion

Cervical cancer is preventable through vaccination against HPV, or screening for cervical precancerous changes which can be treated. Access to cervical precancer treatment in low- and middle-income countries is hindered by a shortage of trained healthcare provider and inadequate health infrastructure. This results in a disproportionately high incidence and mortality rate from this otherwise preventable disease. Use of self-administered topical therapies for cervical precancer treatment, if found to be feasible and effective, can be transformative in increasing access to secondary prevention of cervical cancer for marginalized women globally. Given recent data demonstrating feasibility of topical Artesunate for treatment of HPV-associated anogenital lesions, including vulvar and cervical precancer, it is imperative to understand the pharmacokinetics of intravaginal use to inform studies using this drug in LMICs where malaria is endemic.

## Data Availability

All data to be produced in the present study will be available upon reasonable request to the authors

## Current Status

The study opened for accrual in June 2024.

## Supporting Information Caption

S1 Figure. Schedule of Events.

## Trial registration

The trial is registered under U.S Clinical trial registry (clinicaltrials.gov, NCT06263582).

## List of abbreviations

LMICs: Low- and middle-income countries:
AS: Artesunate
WHO: World Health Organization
CIN2/3: High-grade cervical intraepithelial neoplasia
HPV: Human papillomavirus
DHA: Dihydroartemisinin
PK: Pharmacokinetics
IV: Intravenous
IM: Intramuscular
ACTs: Artemisinin-based combination therapies

## Declarations

### Consent for publication

Not applicable.

### Availability of data and materials

Not applicable.

## Acknowledgments

Artesunate vaginal suppositories (pessaries) were provided as in-kind support by Frantz Viral Therapeutics (Mentor, OH).

## Authors’ contributions

CM conceived and designed the study, providing subject matter expertise and overseeing all aspects of protocol development. JO (Co-Principal Investigator) provided guidance on protocol development and will lead protocol implementation in country. CO (Co-Investigator) contributed to study design, protocol implementation, and capacity building for providers. KS contributed to manuscript writing. GG contributes to lab management activities including collection, storage, and shipment of samples. BM and CC contributed to in-country study coordination activities. LR contributed to study methodology conceptualization, protocol development, and manuscript writing. MP contributed to methodology conceptualization, protocol development, manuscript writing, and implementation. WZ will contribute to formal analysis. All authors, in their respective roles, contributed to study and manuscript preparation and have collectively approved the final manuscript.

